# Difficulty with Mobility among the Aged in Ghana: Evidence from Wave 2 of the World Health Organization’s study on Global Ageing and Adult Health

**DOI:** 10.1101/2023.01.26.23285047

**Authors:** Kingsley Boakye, Antoinette Ama Aidoo, Mohammed Aliyu, Daniel Boateng, Emmanuel Kweku Nakua

**Affiliations:** Department of Epidemiology and Biostatistics, School of Public Health, Kwame Nkrumah University of Science and Technology, Kumasi, Ghana; Julius Global Health, Julius Centre for Health Sciences and Primary Care, University Medical Centre, Utrecht, The Netherlands; St. Patrick’s Nursing and Midwifery Training College, Offinso, Ashanti Region, Ghana

**Keywords:** Difficulty, Mobility, Aged, Elderly, Study on Global Ageing and Adult Health

## Abstract

**Background:** The recent decline in mortality, increase in life expectancy and technological and scientific advancements have led to an increasingly ageing population. About 727 million people globally were aged 65 or more in 2020, and 1 in 6 people will be age 65 years or above by 2050. About 7% of Ghana’s population was over 60 years in 2010, and projected to be 12% by 2050. However, the aged are confronted with degenerative conditions that translate into difficulty with mobility. The study was conducted to investigate the difficulty with mobility among the aged in Ghana.

**Methods:** The study utilised a cross-sectional dataset of the 2014/2015 (wave 2) Study on Global Ageing and Adult Health and included 1,848 participants aged ≥50 years. The survey command was applied to adjust for sampling biases and the design of the study. At 5% alpha level, a chi-square test of independence was conducted to determine the association between dependent and independent variables. At 95% confidence interval and 5% alpha level, three-level multilevel logistic regression models were performed. The fixed-effects were presented in odds ratio and the random effects were presented using the Intra-Class Correlation. All analysis were performed using STATA statistical software version 16.0.

**Results:** Out of the 1,848 participants, 62.3% had difficulty with mobility. Additionally, age (80 and above) [AOR=4.70, 95%CI=2.34 – 9.43], difficulty performing household activities [AOR=6.96, 95%CI=5.03 – 9.64], experiencing bodily pains [AOR=3.21, 95%CI=1.81 – 5.60] and bodily discomfort [AOR=3.39, 95%CI=1.91 – 5.99] and difficulty with vision [AOR=1.70, 95%CI=1.18 – 2.43] had higher odds of difficulty with mobility. However, engaging in vigorous activities [AOR=0.44, 95%CI=0.32 – 0.63] and having good health [AOR=0.41, 95%CI=0.19 – 0.88] were protective of difficulty with mobility.

**Conclusion:** The study concludes that the aged in Ghana had higher prevalence (62.3%) of difficulty with mobility which is associated with age (80 and above), difficulty performing household activities, bodily pains and discomfort, and difficulty with vision. This suggests the need to provide support and assistive devices for the aged and provide geriatric care including recreational fields and care homes to address the health and physical needs of the aged in Ghana.

## Background

The global population is swiftly ageing and this is attributed to significant reductions in mortality at younger ages in low and middle-income countries (LMICs) and continuing increases in life expectancy among the aged globally (Biritwum *et al*., 2013; World Health Organization (WHO), 2015). In the year 2020, an estimated 727 million people were over 65 years and by 2050, an estimated 20% of the global population will be over 65 years (United Nations (UN), 2020). According to WHO, about 2 billion people globally will be over 60 years by 2050 and over 400 million will be aged 80 years and above, out of which 80% will dwell in LMICs (WHO, 2014). This trend in ageing will increase following the scientific, medical and technological advancements globally (Giardini *et al*., 2018).

Ageing is inevitable, however, rapid ageing is problematic for people in LMICs (Tardif, 2014) and this requires critical attention (Porter, Tewodros and Gorman, 2018). Unfortunately, the world is unable to address the swift demands arising from the growing numbers of aged, even in higher-income countries (Tardif, 2014). In sub-Saharan Africa (SSA), including Ghana, the rapid ageing population is presenting challenges to the already weakened health systems. By 2050, SSA is projected to have 8.3% of its population as aged (WHO, 2014). Ghana had 7% of the aged (60+ years) in 2010, a figure among the highest proportions in SSA, and projected to increase to 12% by 2050 (GSS, 2013; Nutakor *et al*., 2021). As of 1960, life expectancy in Ghana was 46.9 years, however, in 2020, it increased to 63.7 years which is further projected to increase to 70 years by 2050, and 76.2 years by 2100 (Awuviry-Newton *et al*., 2021; UN, 2019). Meanwhile, this increase in life expectancy is not tantamount to an increase in good health (Lopes *et al*., 2021). This is a clear-cut indication of the impending burden of care for the aged.

The inadequate preparedness to deal with this demographic shift, coupled with little attention given to the aged is quite problematic (Braimah and Rosenberg, 2021). Ageing brings about a deterioration in health leading to high levels of morbidity and difficulty with mobility, termed as impaired movement (Rosso *et al*., 2011; Braimah & Rosenberg, 2021). Mobility is key to living independently and quality of life. Mobility also accounts for the well-being of the aged population because it promotes healthy living (Cuignet *et al*., 2020). However, mobility limitations are increasingly prevalent among the aged, especially those aged 70 years and above, affecting more than one-third (35%) (Freiberger *et al*., 2020). The difficulty in mobility among the aged contributes to increased falls, hospitalisation, mortality and decreased quality of life (Freiberger *et al*., 2020). Hence, as the population ages, maintaining independent mobility is critical, especially for women who mostly have heightened risk of functional decline and disability (Bergland *et al*., 2017). The difficulty with mobility among aged could result in loss of muscle mass (sarcopenia), osteoporosis and obesity and it is associated with health problems and injuries (Braimah and Rosenberg, 2021). The aged with mobility difficulties have increased rates of morbidity, poorer quality of life and are more probable to be socially isolated (Manini, 2013). Hence, it is important to pay critical attention to the situation of the aged, particularly in areas of research and policy (Braimah and Rosenberg, 2021). There is currently a paucity of literature in Ghana regarding mobility among the aged. This lack of data means there will be inaccurate and unreliable data for policymaking, apt interventions and formulation of policies and programs in relation to mobility (Biritwum *et al*., 2013). The aim of this paper was to investigate difficulty with mobility among the aged in Ghana. Thus, the paper provides current evidence and contributes to bridging the literature gap on difficulty with mobility among the aged in Ghana.

## Methods

### Data source and sampling

This study analysed data from 2014/2015 (Wave 2) Study on Global Ageing and Adult Health (SAGE) (Biritwum *et al*., 2013). In brief, SAGE is a multi-country study that collects data to complement existing ageing data sources to inform policy and programmes. The study employed multi-stage cluster sampling techniques where clusters were systematically sampled with known non-zero selection probability and households residing in the selected clusters identified/listed and individuals in those selected households interviewed for the study The WHO and the University of Ghana Medical School, Department of Community Health collaborated to implement SAGE Wave 2 study. Detailed description of the methods is published elsewhere (Biritwum et al., 2013; Charlton et al., 2016).

The Wave 2 WHO’s SAGE study interviewed 4,735 individuals, primarily focusing on older adults (≥50 years), but for comparison purposes, a smaller sample of those aged 18 to 49 years was also included in the study (Aheto and Dagne, 2021). However, this study was restricted to 1,848 aged (50+ years) who had complete information on variables of interest.

### Dependent variable

The main dependent variable was difficulty with mobility which was computed from a combination of two questions from the questionnaire of the WHO SAGE study; “overall in the last 30 days, how much difficulty did you have with moving around?” and “how much difficulty did you have engaging in vigorous activities?” The questions had responses “none”, “mild”, “moderate”, “severe” and “extreme/cannot do”. The two questions were merged and the response categories, “mild”, “moderate”, “severe” and “extreme/cannot do” recoded as “difficulty”, and “none” was classified as “no difficulty”. “No difficulty” and “difficulty” were then recoded into “0” and “1”, respectively.

### Independent variables

The study considered 18 independent variables to help determine factors associated with difficulty with mobility among the aged in Ghana. The variables were age, place of residence, gender, marital status, ever had formal education, ethnicity, religion, ever worked, perceived health status, difficulty with household activities, bodily pains, bodily discomfort, difficulty with sight/vision, body mass index (BMI), engage in vigorous activities, diagnosed with arthritis, diagnosed with stroke, and diagnosed with depression. These variables have been used to determine difficulty with mobility among the aged elsewhere (McDonald-Miszczak et al., 2001; Shafrin et al., 2017). Additionally, age was recoded into “50 – 59”, “60 – 69”, “70 – 79”, “80 and above; marital status was recoded into “never married”, “married”, “separated/divorced” and “widowed”; ethnicity was recoded into “Akan”, “Ewe”, “Ga-adangbe”, “Gruma/Grusi/Guan” and “Mande-busanga/Mole-dagboni”; religion was recoded “no religion”, “Christian”, “Islam” and “traditionalist”; health status was recoded into “good” and “poor/bad”; difficulty with work was recoded into “no difficulty” and “difficulty”; BMI was recoded into “underweight”, “normal/healthy”, “overweight” and “obese”; difficulty with sight/vision was recoded into “no difficulty” and “difficulty”; bodily discomfort was recoded into “no discomfort” and “discomfort”; diagnosed with depression was recoded into “not depressed” and “depressed”. The initial coding for these selected variables in the WHO’s SAGE study is attached as a supplementary file (Appendix 1).

### Ethical clearance

The SAGE study was approved by the World Health Organization’s Ethical Review Board (reference number RPC149) and the Ethical and Protocol Review Committee, College of Health Sciences, University of Ghana, Accra, Ghana. Written informed consent was obtained from all study respondents. All the SAGE study followed all ethical procedures and methods were performed in accordance with the relevant guidelines and regulations which ensured that participants’ rights were not violated.

### Data Management and analytical procedures

Data for the study was downloaded after authors sought for permission. The data was then cleaned using self-written commands to check for incompleteness and errors in STATA version 16.0. All errors including completeness and consistency were checked before actual analysis.

Data cleaning using self-written commands as well as statistical analysis were performed using STATA statistical software version 16.0 (StataCorp, 2021). First, “survey set command” was applied to account for the sampling biases, complex survey design and generalisability of findings, respectively. Following that, descriptive computations were conducted to describe the general sampled characteristics. At 5% alpha threshold, a chi-square test of independence was conducted to ascertain the association between dependent and independent variables. Collinearity diagnostics was performed and reported using variance inflation factor (VIF). The results indicated no evidence of collinearity between independent variables (Mean VIF=1.65, Maximum VIF=5.05, Minimum VIF=1.02) (see Appendix 2).

A multilevel logistic regression model was conducted to determine the association between the dependent and independent variables. The extension from the single-level model to multilevel regression model is warranted due to the clustering and/or hierarchical structure of the SAGE dataset, where individuals are nested within households/EAs (Aheto and Dagne, 2021). The presence of nesting leads to correlation between observations in each cluster and violation of the independence assumption in Generalised Linear Models (GLM) (Amegbor *et al*., 2020). Hence, the use of a single-level model (GLM) will lead to misspecification and errors in parameter estimates (Heck, 2009; Cofie, 2020) making multilevel the suitable and excellent model. The study considered individuals (ids) as level 1 identifiers and EAs as the level-2 identifier to explain the variability and clustering between and within-groups (Makupe, Kumwenda and Kazembe, 2019).

As such, at 95% confidence interval (95% CI) and 5% alpha threshold, three-stage multilevel regression models were built to assess the relationship between dependent and independent variables. The first was the null model (Model 0) which accounted for the variability of the outcome variables that could be attributed to the clustering of the PSUs/clusters. Subsequently, a second model (Model I) also assessed the crude association between dependent and independent variables. Finally, Model II (adjusted model) was conducted to account for the effect of other covariates. The outputs were presented using Odds Ratio (OR) and 95% CI. Due to the nesting, the Akaike Information Criterion (AIC) was utilised to assess the model fitness and the model with low AIC values was selected as the best model (Portet, 2020).

## Results

Table 1 presents the socio-demographic characteristics of the study participants. Descriptively, more than one-third (39.7%) were aged between 50 – 59 years and nearly one-third (31.9%) were in the category of 60 – 69 years. Little over fifty percent (52.9%) reside in rural areas and were males (56.0%). Nearly two-thirds (60.3%) were married and nearly two-thirds (61.0%) attained formal education. Among those who were educated formally, nearly half (46.5%) had basic education and secondary education (46.1%), respectively. Nearly half of participants (46.9%) were Akans and a little above three-fourths (75.4%) were Christians.

**Table 1:** Socio-demographic characteristics of participants

| Variable | Unweighted |  | Weighted |  |
| --- | --- | --- | --- | --- |
|  | Frequency<br>(n=1,848) | Percentage<br>(%) | Frequency<br>(n=1,848) | Percentage<br>(%) |
| **Age group (years)** |  |  |  |  |
| 50 – 59 | 457 | 24.7 | 457 | 39.7 |
| 60 – 69 | 679 | 36.8 | 679 | 31.9 |
| 70 – 79 | 480 | 26.0 | 480 | 18.9 |
| ≥80 | 232 | 12.5 | 200 | 9.5 |
| **Place of residence** |  |  |  |  |
| Urban | 762 | 41.2 | 762 | 47.1 |
| Rural | 1,086 | 58.8 | 1,086 | 52.9 |
| **Gender** |  |  |  |  |
| Male | 953 | 51.6 | 953 | 56.0 |
| Female | 895 | 48.4 | 895 | 44.0 |
| **Marital status** |  |  |  |  |
| Never married | 62 | 3.3 | 62 | 3.0 |
| Married | 981 | 53.1 | 981 | 60.3 |
| Separated/divorced | 234 | 12.7 | 234 | 11.8 |
| Widowed | 571 | 30.9 | 571 | 24.9 |
| **Ever had formal education** |  |  |  |  |
| No | 880 | 47.6 | 880 | 39.0 |
| Yes | 968 | 52.4 | 968 | 61.0 |
| **If yes, highest educational level (n=968)** |  |  |  |  |
| Basic | 429 | 44.3 | 429 | 46.5 |
| Secondary | 470 | 48.6 | 470 | 46.4 |
| Tertiary | 69 | 7.1 | 69 | 7.1 |
| **Ethnicity** |  |  |  |  |
| Akan | 932 | 50.4 | 932 | 46.9 |
| Ewe | 130 | 7.0 | 130 | 11.0 |
| Ga-adangbe | 214 | 11.6 | 214 | 13.1 |
| Gruma/Grusi/Guan | 129 | 7.0 | 129 | 6.2 |
| Mande-busanga/mole-dabani | 443 | 22.8 | 443 | 22.8 |
| **Religion** |  |  |  |  |
| Christian | 1,354 | 73.3 | 1,354 | 75.4 |
| Moslem | 327 | 17.7 | 327 | 17.1 |
| Traditionalist | 105 | 5.7 | 105 | 4.5 |
| No religion | 62 | 3.3 | 62 | 3.0 |

### Difficulty with mobility among the aged in Ghana

Figure 1 is a pictorial presentation of the weighted prevalence of difficulty with mobility among the aged in Ghana. Nearly two-thirds of participants (62.3%) had difficulty with mobility and more than one-third (37.7%) had no difficulty.

**Figure 1:**
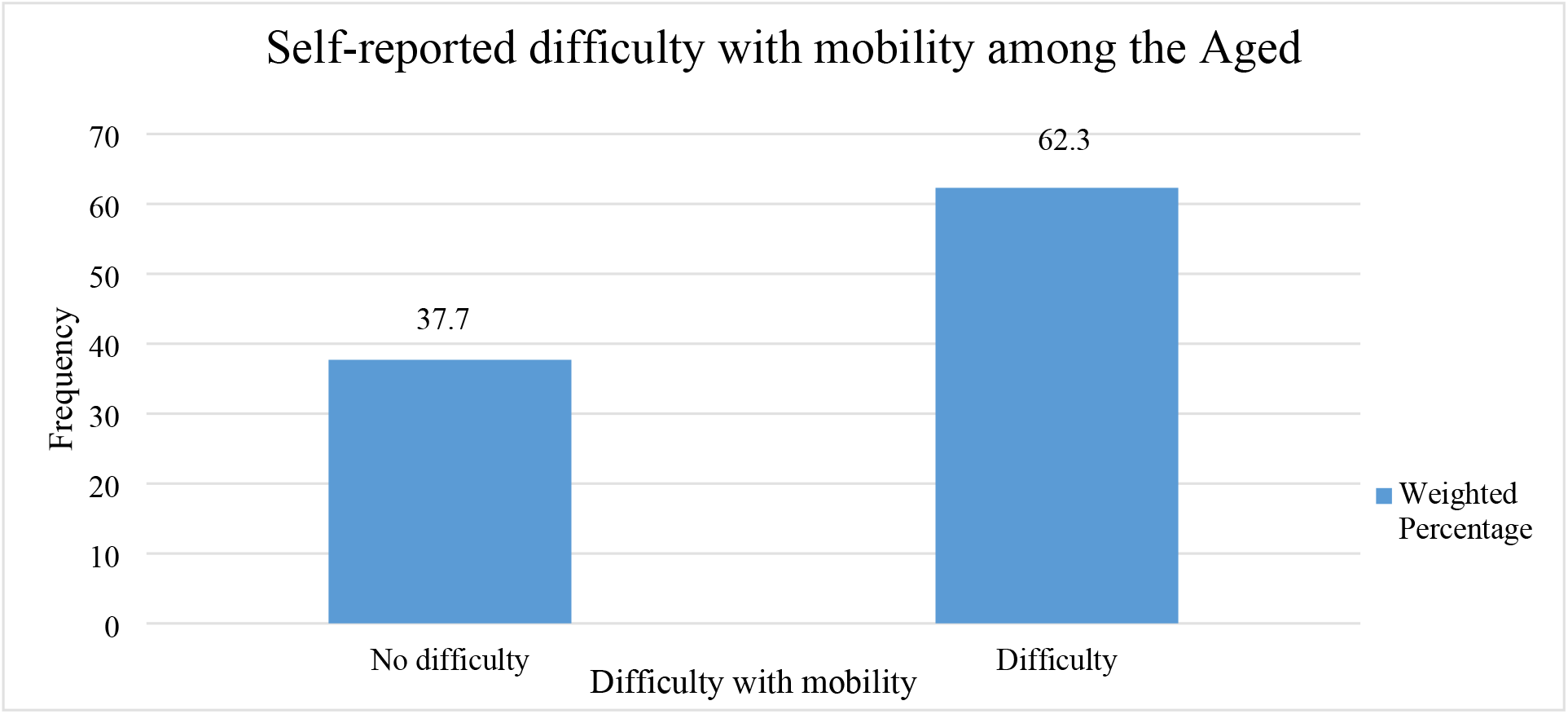
Self-reported difficulty with mobility among the aged in Ghana.

The results of the chi-square test of independence showed that age group, gender, marital status, ever attended school, health status, difficulty with work/house activities, experience bodily pains, bodily discomfort, visual difficulty, BMI, engage in vigorous activities and diagnosed of depression were associated with difficulty with mobility (Table 2).

**Table 2:**
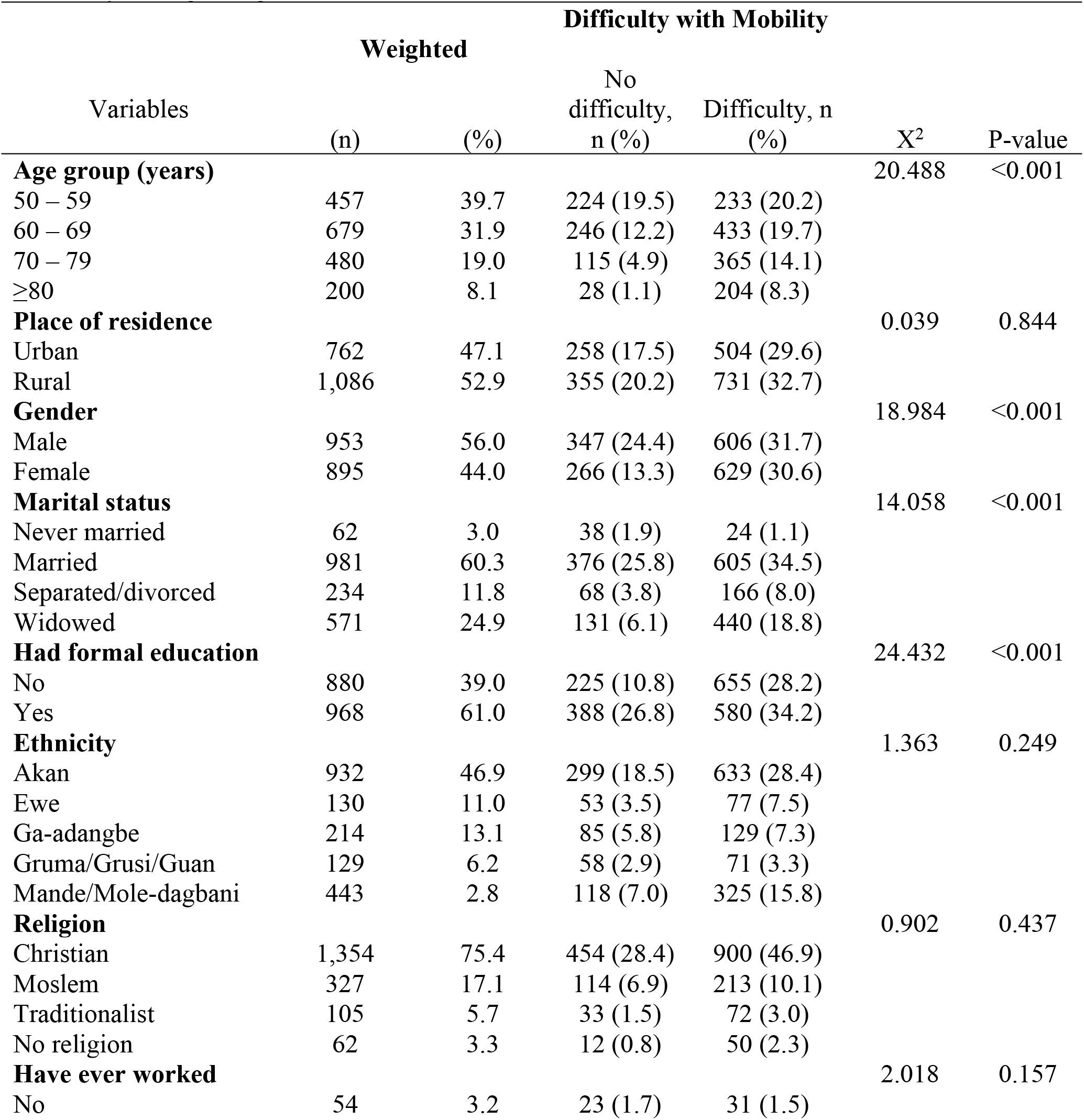
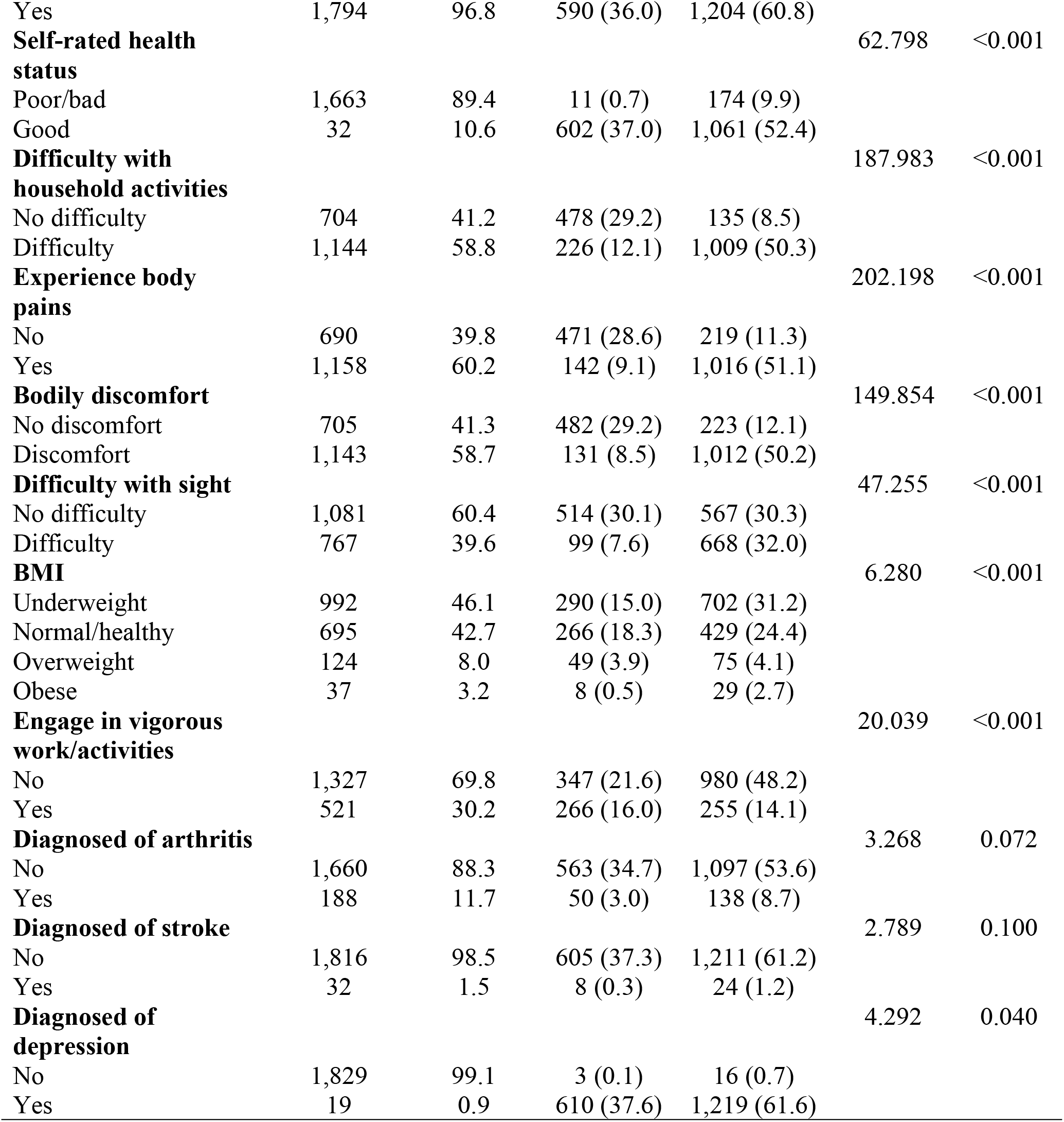
Bivariate analysis of demographic features and factors associated with difficulty with mobility among the aged

| Variables | Weighted |  | Difficulty with Mobility |  | X<sup>2</sup> | P-value |
| --- | --- | --- | --- | --- | --- | --- |
|  | (n) | (%) | No difficulty, n (%) | Difficulty, n (%) |  |  |
| **Age group (years)** |  |  |  |  | 20.488 | <0.001 |
| 50 – 59 | 457 | 39.7 | 224 (19.5) | 233 (20.2) |  |  |
| 60 – 69 | 679 | 31.9 | 246 (12.2) | 433 (19.7) |  |  |
| 70 – 79 | 480 | 19.0 | 115 (4.9) | 365 (14.1) |  |  |
| ≥80 | 200 | 8.1 | 28 (1.1) | 204 (8.3) |  |  |
| **Place of residence** |  |  |  |  | 0.039 | 0.844 |
| Urban | 762 | 47.1 | 258 (17.5) | 504 (29.6) |  |  |
| Rural | 1,086 | 52.9 | 355 (20.2) | 731 (32.7) |  |  |
| **Gender** |  |  |  |  | 18.984 | <0.001 |
| Male | 953 | 56.0 | 347 (24.4) | 606 (31.7) |  |  |
| Female | 895 | 44.0 | 266 (13.3) | 629 (30.6) |  |  |
| **Marital status** |  |  |  |  | 14.058 | <0.001 |
| Never married | 62 | 3.0 | 38 (1.9) | 24 (1.1) |  |  |
| Married | 981 | 60.3 | 376 (25.8) | 605 (34.5) |  |  |
| Separated/divorced | 234 | 11.8 | 68 (3.8) | 166 (8.0) |  |  |
| Widowed | 571 | 24.9 | 131 (6.1) | 440 (18.8) |  |  |
| **Had formal education** |  |  |  |  | 24.432 | <0.001 |
| No | 880 | 39.0 | 225 (10.8) | 655 (28.2) |  |  |
| Yes | 968 | 61.0 | 388 (26.8) | 580 (34.2) |  |  |
| **Ethnicity** |  |  |  |  | 1.363 | 0.249 |
| Akan | 932 | 46.9 | 299 (18.5) | 633 (28.4) |  |  |
| Ewe | 130 | 11.0 | 53 (3.5) | 77 (7.5) |  |  |
| Ga-adangbe | 214 | 13.1 | 85 (5.8) | 129 (7.3) |  |  |
| Gruma/Grusi/Guan | 129 | 6.2 | 58 (2.9) | 71 (3.3) |  |  |
| Mande/Mole-dagbani | 443 | 2.8 | 118 (7.0) | 325 (15.8) |  |  |
| **Religion** |  |  |  |  | 0.902 | 0.437 |
| Christian | 1,354 | 75.4 | 454 (28.4) | 900 (46.9) |  |  |
| Moslem | 327 | 17.1 | 114 (6.9) | 213 (10.1) |  |  |
| Traditionalist | 105 | 5.7 | 33 (1.5) | 72 (3.0) |  |  |
| No religion | 62 | 3.3 | 12 (0.8) | 50 (2.3) |  |  |
| **Have ever worked** |  |  |  |  | 2.018 | 0.157 |
| No | 54 | 3.2 | 23 (1.7) | 31 (1.5) |  |  |
| Yes | 1,794 | 96.8 | 590 (36.0) | 1,204 (60.8) |  |  |
| **Self-rated health status** |  |  |  |  | 62.798 | <0.001 |
| Poor/bad | 1,663 | 89.4 | 11 (0.7) | 174 (9.9) |  |  |
| Good | 32 | 10.6 | 602 (37.0) | 1,061 (52.4) |  |  |
| **Difficulty with household activities** |  |  |  |  | 187.983 | <0.001 |
| No difficulty | 704 | 41.2 | 478 (29.2) | 135 (8.5) |  |  |
| Difficulty | 1,144 | 58.8 | 226 (12.1) | 1,009 (50.3) |  |  |
| **Experience body pains** |  |  |  |  | 202.198 | <0.001 |
| No | 690 | 39.8 | 471 (28.6) | 219 (11.3) |  |  |
| Yes | 1,158 | 60.2 | 142 (9.1) | 1,016 (51.1) |  |  |
| **Bodily discomfort** |  |  |  |  | 149.854 | <0.001 |
| No discomfort | 705 | 41.3 | 482 (29.2) | 223 (12.1) |  |  |
| Discomfort | 1,143 | 58.7 | 131 (8.5) | 1,012 (50.2) |  |  |
| **Difficulty with sight** |  |  |  |  | 47.255 | <0.001 |
| No difficulty | 1,081 | 60.4 | 514 (30.1) | 567 (30.3) |  |  |
| Difficulty | 767 | 39.6 | 99 (7.6) | 668 (32.0) |  |  |
| **BMI** |  |  |  |  | 6.280 | <0.001 |
| Underweight | 992 | 46.1 | 290 (15.0) | 702 (31.2) |  |  |
| Normal/healthy | 695 | 42.7 | 266 (18.3) | 429 (24.4) |  |  |
| Overweight | 124 | 8.0 | 49 (3.9) | 75 (4.1) |  |  |
| Obese | 37 | 3.2 | 8 (0.5) | 29 (2.7) |  |  |
| **Engage in vigorous work/activities** |  |  |  |  | 20.039 | <0.001 |
| No | 1,327 | 69.8 | 347 (21.6) | 980 (48.2) |  |  |
| Yes | 521 | 30.2 | 266 (16.0) | 255 (14.1) |  |  |
| **Diagnosed of arthritis** |  |  |  |  | 3.268 | 0.072 |
| No | 1,660 | 88.3 | 563 (34.7) | 1,097 (53.6) |  |  |
| Yes | 188 | 11.7 | 50 (3.0) | 138 (8.7) |  |  |
| **Diagnosed of stroke** |  |  |  |  | 2.789 | 0.100 |
| No | 1,816 | 98.5 | 605 (37.3) | 1,211 (61.2) |  |  |
| Yes | 32 | 1.5 | 8 (0.3) | 24 (1.2) |  |  |
| **Diagnosed of depression** |  |  |  |  | 4.292 | 0.040 |
| No | 1,829 | 99.1 | 3 (0.1) | 16 (0.7) |  |  |
| Yes | 19 | 0.9 | 610 (37.6) | 1,219 (61.6) |  |  |

Table 3 presents the multi-level logistic regression results of difficulty with mobility among the aged. Participants aged 80 and above compared to 50 – 59 years [OR=13.05, 95%CI=7.50 – 21.95], females compared to males [OR=1.54, 95%CI=1.22 – 1.95], widows compared to never married [OR=6.20, 95%CI=3.24 – 11.8] and those with difficulty with household activities [OR=17.9, 95%CI=13.6 – 23.6] had higher odds of difficulty with mobility. Additionally, participants who experienced bodily pains compared to no pains [OR=17.0, 95%CI=13.0 – 22.3], experienced bodily discomfort compared to no discomfort [OR=20.0, 95%CI=15.0 – 26.4] and had difficulty with sight compared with no difficulty [OR=6.31, 95%CI=4.78 – 8.33] had higher odds of difficulty with mobility. Moreover, respondents who had formal education compared to no formal education [OR=0.40, 95%CI=0.31 – 0.52], self-reported to have good health compared to poor health [OR=0.08, 95%CI=0.04 – 0.15], those with normal weight compared to underweight [OR=0.65, 95%CI=0.51 – 0.83], those who engaged in vigorous activities compared to those who do not [OR=0.24, 95%CI=0.18 – 0.31] and those diagnosed with depression compared to those who are not [OR=0.23, 95%CI=0.06 – 0.92] had decreased odds of difficulty with mobility.

**Table 3:**
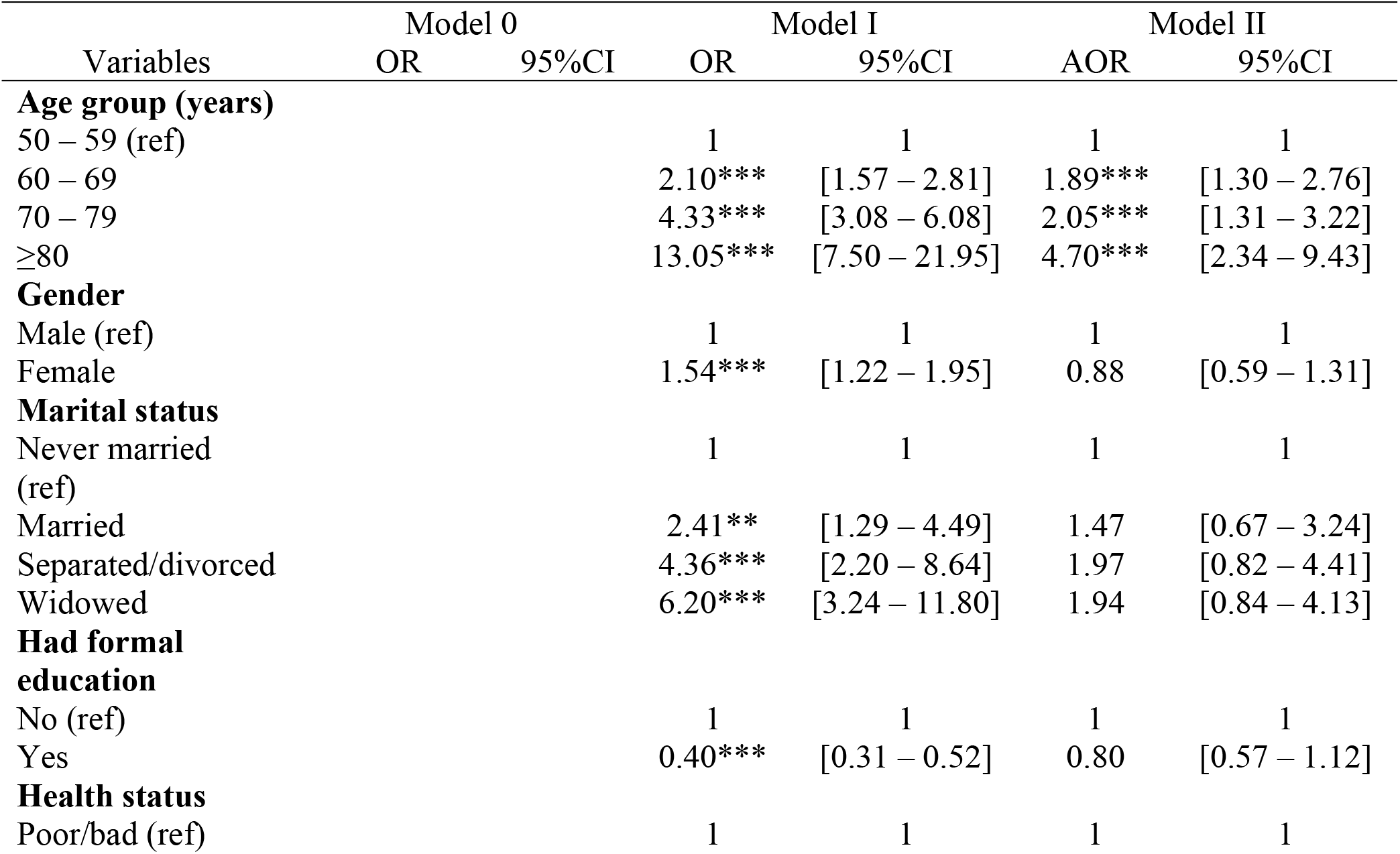
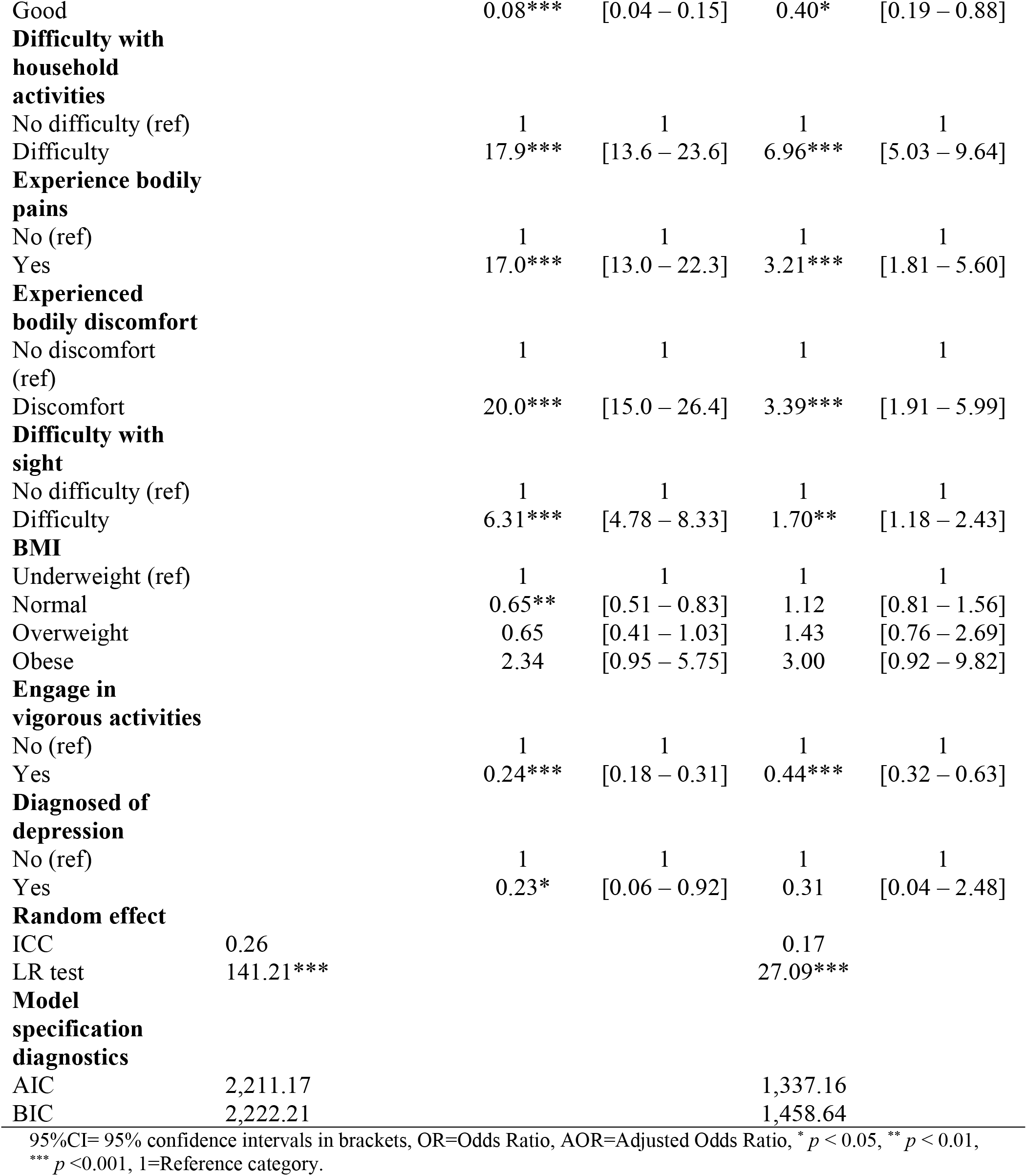
Multilevel mixed-effects logistic regression results of difficulty with mobility

In the adjusted model (Model II), participants aged 80 and above compared to 50 – 59 years [AOR=4.70, 95%CI=2.34 – 9.43], participants with difficulty performing household activities compared to no difficulty [AOR=6.96, 95%CI=5.03 – 9.64] participants who experienced bodily pains compared to no pains [AOR=3.21, 95%CI=1.81 – 5.60], and experienced bodily discomfort compared to no discomfort [AOR=3.39, 95%CI=1.91 – 5.99] had higher odds of difficulty with mobility.

## Discussion

We investigated the difficulty with mobility among the aged (≥50 years) in Ghana. The key findings were that among the aged (≥50 years) in Ghana, nearly two-thirds (62.3%) had difficulty with mobility. Moreover, difficulty with mobility among the aged in Ghana is associated with age (80 and above), difficulty performing household activities, bodily pains and discomfort, and difficulty with vision.

The study revealed that nearly two-thirds (62.3%) of the aged had difficulty with mobility. In congruence to the study’s findings, a global study by Webber et al. (2010) revealed that about half of people aged 65 years or older reported difficulties with respect to walking or climbing stairs (Webber, Porter and Menec, 2010). Other studies conducted among the aged in other parts of Africa however reported lesser prevalence. For instance, a study conducted among the aged (≥50 years) in Nigeria revealed that 35.8% had difficulty with mobility (Balogun and Guntupalli, 2016) indicating that nearly two-thirds (64.2%) had no difficulty with mobility. Also, Yaya and colleagues had a contrasting finding in their cross-national study in South Africa and Uganda. The study revealed that 20% of the aged had difficulty with mobility (Yaya *et al*., 2020). The likely justification for the difficulty with mobility is the loss of strength and function that is characterised by sarcopenia in adulthood (Billot *et al*., 2020) leading to slow walking, less stability, inefficiency, poor timing and coordination of postures and poor gaiting (Brach and Vanswearigen, 2013). Another plausible explanation is the alteration of muscular strength during old age (Billot *et al*., 2020) and decrease in physical strength each year after sixty years (Grimmer *et al*., 2019; Billot *et al*., 2020). Despite the high prevalence of difficulty with mobility reported across studies, variations exist in the prevalence estimates which could be attributed to the differences in study settings, genetic make-up and study designs employed.

This study revealed that participants aged 80 years and above had increased odds of difficulty with mobility. It is logical to argue that mobility difficulties increases with ageing stemming from the deterioration in functioning and degenerative conditions owing to sarcopenia (Khan, 2018). This argument is supported by research in Ghana that revealed that the aged are mostly confronted with degenerative conditions and physical deterioration that affect their capacity to effectively function including mobility (Kpessa-Whyte and Tsekpo, 2020). Even though, mobility challenges however could be prevalent at younger ages, research revealed that mobility difficulties are mostly prevalent at old age which could also be attributed to the decline in strength in the muscles (Miszkurka *et al*., 2012). A research conducted in Nigeria also revealed that the aged (>70 years) are at heightened risk of mobility difficulties (Balogun and Guntupalli, 2016). This decline in mobility could be present even in the absence of co-morbidity because ageing induces biological and functional decline at several levels (including loss of muscle strength and mass and decline in balance) resulting in difficulty in mobility (Billot *et al*., 2020).

This study further revealed that participants who engage in vigorous activities or exercise had lower odds of difficulty with mobility and participants with difficulty with mobility had difficulty exercising or performing household activities even though, reverse causality cannot be completely ruled out in this association. Corroborating and affirming the finding, a study in Canada concluded that the aged who engage in vigorous activities have reduced risk of difficulty with mobility (Paterson and Warburton, 2010). Similarly, in Taiwan, research revealed that engaging in household activities was associated lower odds of mobility limitations (Lêng and Wang, 2016). This is because, engaging in household activities and exercise make the aged physically active and be more productive thereby increasing mobility (Lêng and Wang, 2016). Researchers and scholars have chronicled that not engaging in vigorous activities or exercise is an independent determinant for mobility decline among the aged (Daley and Spinks, 2000). The plausible explanation is that engagement in exercise or physical and household activities help in contracting the skeletal muscle leading to an increase in energy expenditure thereby reducing risk for mobility limitations (Welmer *et al*., 2013). Another justifiable reason is that these activities are characterized by planned, structured, and repetitive movement which can preserve or improve physical determinants of mobility thereby reducing the odds of difficulty with mobility or otherwise.

The study also revealed that participants who experienced bodily discomfort and pains had increased odds of difficulty with mobility. In a related study in West Africa, health conditions including bodily pains and discomfort are risk factors for mobility disability (Miszkurka *et al*., 2012). Consistent with the above finding, in USA, research found that the presence of mild-to-moderate pain among the aged was independently associated with poor or difficulty with mobility among the aged (Schepker *et al*., 2017). The association between pain and mobility limitation could however be as a result of reverse causation. Musich et al. for instance, in their study in the USA found that higher levels of mobility limitations were linked to pain among the aged. The authors, therefore, recommended mobility-enhancing interventions that could promote successful ageing (Musich *et al*., 2018).

Participants who had difficulty with vision had higher odds of difficulty with mobility. Similarly, in a study in rural India, the authors found mobility losses of 5.1, 10.2, and 23.4 points of 100 among older people with visual difficulties, low vision, and blindness, respectively (Nirmalan *et al*., 2005). This finding is also similar to other related studies conducted in high-income countries. For instance, related studies in Israel, Canada, China, and USA that found that the aged who had visual difficulty had walking and stair climbing difficulties (Jacobs et al., 2005; Popescu et al., 2011; Fenwick et al., 2016; Swenor et al., 2015). Research revealed that older adults with vision difficulties had significantly difficulty with mobility including slower walking speeds compared to non-visually impairment individuals (Miyata *et al*., 2021). The likely explanation is that the slowness in walking speed is a way to maintain or improve their mobility safety.

Additionally, the study revealed that the aged who reported having good health status compared with poor health had decreased odds of difficulty with mobility. Poor health has been shown to be a determinant of mobility limitations among the aged in a similar study in Nigeria (Balogun and Guntupalli, 2016). In other words, the aged who perceived to have good health (that is, devoid of cardiovascular and metabolic diseases, osteoporosis, muscular weakness) had decreased odds of difficulty with mobility (McPhee *et al*., 2016). In congruence to the above, a study published elsewhere revealed that difficulties with mobility is central to healthy ageing (Satariano *et al*., 2012). The authors further justified that having poor health is associated with limitations in both walking and driving. Interestingly, research in the United Kingdom revealed that even older adults who perceived their health to be good are not devoid of mobility limitations due to ageing (Degens *et al*., 2021). However, among these healthy older people, mobility limitation is due to weakness of the body and not slower muscle contractile properties (Degens *et al*., 2021).

### Strength and limitations

The study is novel because it presented updated information on difficulty with mobility among the aged in Ghana. Also, the study utilised secondary data from nationally representative cross-sectional survey with relatively large sample size (n=1,848). Additionally, the researcher applied rigorous, advanced statistical and analytical methods to analyse the data for the study making it robust. However, despite the above-mentioned strengths, the study had its limitations that cannot be overemphasised. The study utilised a dataset that was collected from respondents’ self-report making recall bias inevitable. The study is also liable to social desirability biases (tendency of respondents to bias responses to make it appropriate or socially acceptable) due to the cross-sectional nature of SAGE study. The cross-sectional nature of the survey also led to the failure to establish a causal relationship. Furthermore, some of the associations found in this study could be due to reverse causality.

### Conclusion

The aged in Ghana had higher prevalence (62.3%) of difficulty with mobility which is associated with age (80 and above), difficulty performing household activities, bodily pains and discomfort, and difficulty with vision. This suggests the need to provide geriatric care including assistive devices, care homes and recreational fields to address the health and physical needs of the aged in Ghana.

## Data Availability

The datasets generated and/or analysed during this current study are freely available upon making official request to WHO-SAGE Team through the WHO website at http://www.who.int/healthinfo/sage/cohorts/en/. However, registration is required for access to the data files.

http://www.who.int/health-info/sage/cohorts/en/

## DECLARATION

### Ethics approval and consent to participate

The authors of this manuscript did not participate in the actual data gathering processes. Hence, we sought permission by officially writing for the dataset. However, the WHO’s Wave 2 study sought ethical approval from the World Health Organization’s Ethical Review Board (reference number RPC149) and the Ethical and Protocol Review Committee, College of Health Sciences, University of Ghana, Accra, Ghana. The SAGE study followed all ethical procedures and ensured that participants’ rights were not violated. Written informed consent was obtained from all study respondents and the dataset was anonymised before making it available to the public.

### Consent for publication

Not applicable

### Competing interests

The authors declare no competing interest. The views expressed in this paper are those of the authors. No official endorsement by the World Health Organization or Ministry of Health of Ghana/Ghana Health Service is intended or should be inferred.

### Funding

No funding was received for the study

### Author’s contributions

KB, DB, and EKN developed the concept. KB conducted the formal analysis and interpreted the results. KB, AAA, MA, DB, and EKN drafted the manuscript. All authors proofread the manuscript for important intellectual content.

## Acknowledgements

For the SAGE, financial support was provided by the U.S. National Institute on Aging through Interagency Agreements (OGHA 04034785; YA1323-08-CN-0020; Y1-AG-1005-01) with the World Health Organization and a Research Project Grant (R01 AG034479-64401A1). WHO contributed financial and human resources to SAGE. The Ministry of Health, Ghana, is supportive of SAGE. The University of Ghana’s Department of Community Health contributed training facilities, data entry support, and storage of materials. The Ghana Statistical Office provided the sampling information for the sampling frame and updates.

## Notes

### Competing Interest Statement

The authors have declared no competing interest.

### Funding Statement

The authors received no specific funding for this work

### Author Declarations

World Health Organization's Ethical Review Board (reference number RPC149) and the Ethical and Protocol Review Committee, College of Health Sciences, University of Ghana, Accra, Ghana.

